# Impact of Vaccine Prioritization Strategies on Mitigating COVID-19: An Agent-Based Simulation Study using an Urban Region in the United States

**DOI:** 10.1101/2021.03.12.21253447

**Authors:** Hanisha Tatapudi, Rachita Das, Tapas K. Das

## Abstract

**Background:** Approval of novel vaccines for COVID-19 had brought hope and expectations, but not without additional challenges. One central challenge was understanding how to appropriately prioritize the use of limited supply of vaccines. This study examined the efficacy of the various vaccine prioritization strategies using the vaccination campaign underway in the U.S.

**Methods:** The study developed a granular agent-based simulation model for mimicking community spread of COVID-19 under various social interventions including full and partial closures, isolation and quarantine, use of face mask and contact tracing, and vaccination. The model was populated with parameters of disease natural history, as well as demographic and societal data for an urban community in the U.S. with 2.8 million residents. The model tracks daily numbers of infected, hospitalized, and deaths for all census age-groups. The model was calibrated using parameters for viral transmission and level of community circulation of individuals. Published data from the Florida COVID-19 dashboard was used to validate the model. Vaccination strategies were compared using a hypothesis test for pairwise comparisons.

**Results:** Three prioritization strategies were examined: a minor variant of CDC’s recommendation, an age-stratified strategy, and a random strategy. The impact of vaccination was also contrasted with a no vaccination scenario. The study showed that the campaign against COVID-19 in the U.S. using vaccines developed by Pfizer/BioNTech and Moderna 1) reduced the cumulative number of infections by 10% and 2) helped the pandemic to subside below a small threshold of 100 daily new reported cases sooner by approximately a month when compared to no vaccination. A comparison of the prioritization strategies showed no significant difference in their impacts on pandemic mitigation.

**Conclusions:** Even though vaccines for COVID-19 were developed and approved much quicker than ever before, their impact on pandemic mitigation was small as the explosive spread of the virus had already infected a significant portion of the population, thus reducing the susceptible pool. A notable observation from the study is that instead of adhering strictly to a sequential prioritizing strategy, focus should perhaps be on distributing the vaccines among all eligible as quickly as possible, after providing for the most vulnerable. As much of the population worldwide is yet to be vaccinated, results from this study should aid public health decision makers in effectively allocating their limited vaccine supplies.

## INTRODUCTION

SARS-CoV-2 and resulting COVID-19 disease has been raging world-wide since early 2020, killing over 2.0 million globally and nearly 450,000 in the United States by the end of January 2021 [1]. A significant winter swell in cases was underway in the U.S. between November 2020 and January 2021 despite protective measures in place such as face mask usage, limited contact tracing, travel restrictions, social distancing practices, and partial community closures. To combat this, many promising novel vaccines were developed, of which two (Pfizer/BioNTech and Moderna) were authorized for emergency use in mid-December 2020 by the U.S. Food and Drug Administration (USFDA) [2]. Data from initial trials of cohorts greater than 30,000 people showed that these vaccines, given in two doses, were safe and have ∼95% effectiveness in preventing COVID-19 [3]. Vaccine deployment in the U.S. began soon after USFDA approval.

Implementing an effective vaccination campaign was essential to dramatically reduce the infection, hospitalization, and death rates, but posed many unique challenges. Vaccine prioritization and allocation strategy was at the forefront of the challenges to effectively vaccinate communities. Strategy was influenced by a number of key factors: 1) limited initial vaccine supply in the months following release, 2) transmission and severity of COVID-19 varying by segment of the population, 3) vaccine approvals only for adults, and 4) acceptability and compliance in the community for two dose vaccination [4].

U.S. Centers for Disease Control (CDC) released an outline prioritizing healthcare personnel, first responders, persons with high risk medical conditions for COVID-19, and older adults >65 years. These groups were given priority for vaccination in phase 1, when supply was limited. In phase 2 (supply increased to begin to meet demand) and phase 3 (supply was greater than demand), other population groups were vaccinated based on age and availability [5]. Vaccine allocation structures with basic similarities and some key differences were used by countries around the world. For example, after healthcare workers, France’s vaccine allocation scheduled other general workers regardless of age who they had determined to be at high risk of contracting and spreading the virus due to contact with the general public. This includes retail, school, transportation, and hospitality staff [6]. Such differences in vaccine prioritization strategies were untested at the time of the study and warranted modeling and examination.

The goal of this paper was to investigate the impact of vaccination on the pandemic via outcome measures such as numbers of infected, hospitalized, and deaths in the months following December 15, 2020 when vaccination would begin in the U.S. Two specific objectives of our investigation were: 1) to assess the expected impact of the vaccination program on mitigating COVID-19, and 2) to inform public health officials on the comparative benefits, if any, of the different vaccine prioritization strategies. We conducted our investigation by using our agent-based (AB) simulation model for COVID-19 that was presented during the early phase of the pandemic [7]. For this study, we first extended calibration of our model till December 30, 2020 to ensure that our model appropriately tracked the explosive increase in cases that started with the onset of winter and the year-end holiday period in 2020. We then enhanced the AB simulation model by adding a framework for vaccination. This included: vaccination priorities for people based on attributes including profession and age, use of two different vaccines by Pfizer/BioNTech and Moderna with their contracted quantities and approximate delivery timelines, vaccine acceptance, transition period between each priority group, vaccination rate, and immunity growth for vaccinated starting with the first dose.

As in [7], we implemented our calibrated AB model, augmented with vaccination, for Miami-Dade County of the U.S. with 2.8 million population, which had been an epicenter of COVID-19 in the U.S. We conducted our investigation by implementing a number of prioritization strategies and obtaining the corresponding numbers of total infections, reported infections, hospitalizations, and deaths. The strategies implemented were 1) minor variant of the CDC recommended strategy, 2) age stratified strategy, and 3) random strategy. We also implemented a no vaccination case. These strategies are explained in a later section. We compared and contrasted the results to assess vaccination efficacy and relative performances of the priority strategies. We made a number of key observations from the results, which we believe will help public health officials around the world to choose effective vaccine prioritization strategies to mitigate the negative impacts of COVID-19.

## LITERATURE REVIEW

On a global scale, equitable and ethical distribution of vaccines for all (low, medium, and high-income) countries is an important question. As the world leader in promoting global health, World Health Organization (WHO) released an evidence-based framework for vaccine-specific recommendations [8]. WHO proposed vaccine prioritization for three potential scenarios of transmission: community transmission, sporadic cases or cluster of cases, and no cases. Each scenario has three stages and focuses on different risk groups. COVID-19 pandemic resembles “community transmission.” For this, the first stage focused on healthcare workers and older adults with highest risk; second stage continued the focus on older adults and people with comorbidities, sociodemographic groups, and educational staff; and the third stage focused on essential workers and social/employment groups unable to physically distance themselves.

National Academy of Sciences, Engineering, and Medicine (NAESM) developed a more comprehensive phased framework for equitable allocation of COVID-19 vaccine [9]. The first phase prioritized healthcare workers and first responders, people with high risk comorbidities, and older adults in crowded living conditions; second phase focused on K-12 school staff and child care workers, essential workers, people with moderate risk comorbidities, people living in shelters, physically and mentally disabled people and staff that provide care, employment settings where social distancing was not possible, and remaining older adults; third phase prioritized young adults, children, and workers; and fourth phase included everyone else. No specific studies had been presented to the literature at the time of this research that had evaluated the efficacy of the proposed vaccination priorities for mitigating COVID-19.

A number of studies can be found in the literature on vaccination strategies for controlling outbreaks of other viruses. The work presented in [10] analyzes the effect of both CDC guided targeted vaccination strategy as well as a mass vaccination strategy for seasonal Influenza outbreaks in the U.S. The study found that a mass vaccination policy reaped the most benefits both in terms of cost and quality-of-life years (QALYs) lost. Authors in [11] use a genetic algorithm to find optimal vaccine distribution strategies that minimize illness and death for Influenza pandemics with age specific attack rates similar to the 1957–1958 A(H2N2) Asian Influenza pandemic and the 1968–1969 A(H3N2) Hong Kong Influenza pandemic. They consider coverage percentage under varying vaccine availability and developed an optimal vaccination approach that was 84% more effective than random vaccination. A study reported in [12] examined vaccination to prevent interpandemic Influenza for high-risk groups and children, and recommended concentrating on schoolchildren, who were most responsible for transmission, and then extended to high-risk groups. A compartmental model in [13] was used to develop optimal strategies to reduce the morbidity and mortality of the H1N1 pandemic. The study found that age specific vaccination schedules had the most beneficial impact on mortality.

It can be concluded from the above review of relevant literature that there is no ‘one size fits all’ strategy for vaccination to either prevent a pandemic outbreak or mitigate one. Virus epidemiology and corresponding disease characteristics, as well as the efficacy and supply of the vaccine must be considered in developing an effective vaccination prioritization strategy. Our paper aims to address this need by presenting a detailed AB simulation modeling approach and using it to assess efficacy of vaccine prioritization strategies for COVID-19.

## METHODOLOGY

Published COVID-19 modeling approaches were either data-driven models, as in [14]–[18], or variants of SEIR type compartmental models as in [19]–[23]. Data driven models are very well suited for understanding the past progression of a pandemic and also for estimating parameters characterizing virus epidemiology. However, these models offer limited ability to predict the future progression of a pandemic that could be dynamically evolving with regards to virus epidemiology, disease manifestations, and sociological conditions. Compartmental models, on the other hand, are aggregate in nature and do not adapt well to changing dynamics of disease transmission. An AB modeling approach is considered to be more suitable for a detailed accounting of individual attributes, specific disease natural history, and complex social interventions [24]. Hence we adopted the AB model approach to conduct our study.

The AB simulation-based methodology was particularized using data for Miami Dade County of Florida, with 2.8 million population, an epicenter for COVID-19 spread in the South-Eastern United States. A step by step approach for building such a model for another region can be found in [7]. The methodology began by generating the individual people according to the U.S. census data that gives population attributes including age (see Table A1 in [7]) and occupational distribution (see Table A4 in [7]). Thereafter, it generated the households based on their composition characterized by the number of adults and children (see Table A2 in [7]). The model also generated, per census data, schools (see Table A3 in [7]) and the workplaces and other community locations (see Table A4 in [7]). Each person was assigned a workplace and household based on the numbers, sizes, and compositions of households, schools and workplaces derived from census data (see Tables A2, A3, and A4 in [7] for distribution of household, schools, and workplaces, respectively). A daily (hour by hour) schedule was assigned to every individual, chosen from a set of alternative schedules based on their attributes. The schedules vary between weekdays and weekends and also depend on the prevailing social intervention orders (see Table A1 in Additional file 1). The methodology incorporated means to implement a number of intervention orders including full and partial closures/reopening of schools and workplaces [7], [25], isolation and quarantine of infected individuals and household members, use of face mask, contact tracing of asymptomatic and pre-symptomatic individuals, and vaccination priorities. The timeline for interventions implemented in the model are summarized in Table 1.

**Table 1:**
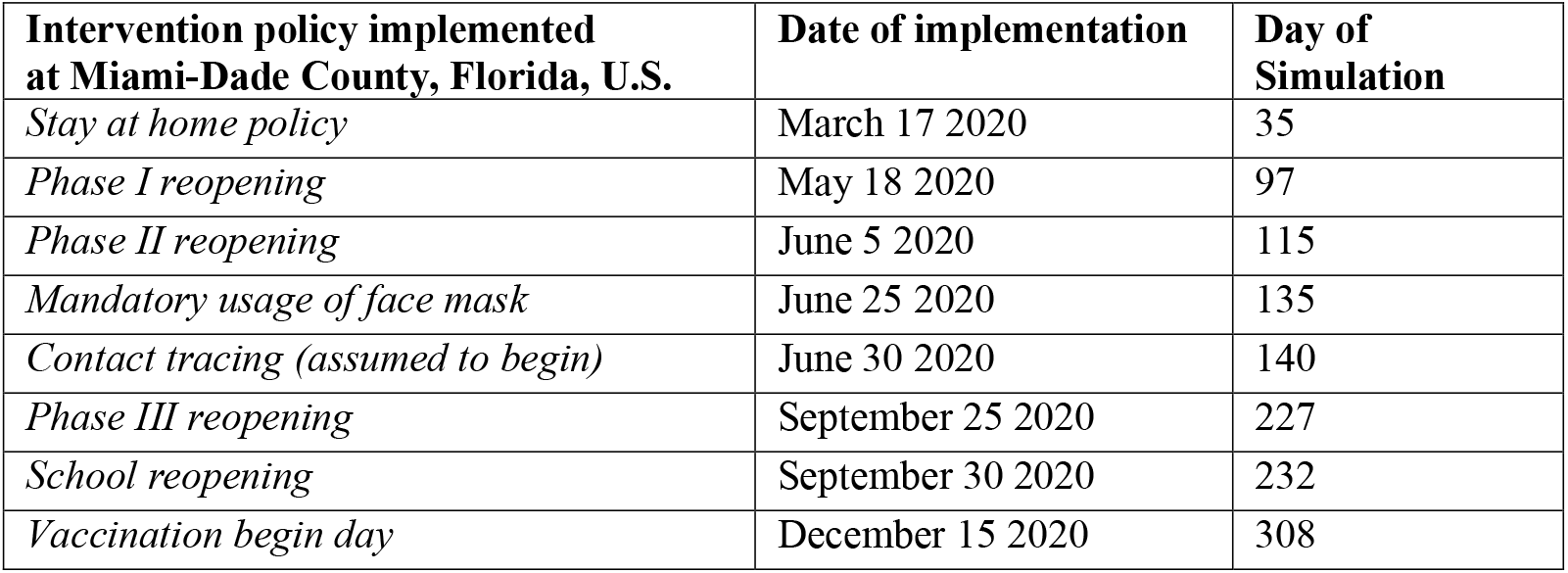
Social intervention order timeline for Miami-Dade County

| **Intervention policy implemented at Miami-Dade County, Florida, U.S.** | **Date of implementation** | **Day of Simulation** |
| --- | --- | --- |
| *Stay at home policy* | March 17 2020 | 35 |
| *Phase I reopening* | May 18 2020 | 97 |
| *Phase II reopening* | June 5 2020 | 115 |
| *Mandatory usage of face mask* | June 25 2020 | 135 |
| *Contact tracing (assumed to begin)* | June 30 2020 | 140 |
| *Phase III reopening* | September 25 2020 | 227 |
| *School reopening* | September 30 2020 | 232 |
| *Vaccination begin day* | December 15 2020 | 308 |

The AB model also included a number of uncertainties such as 1) time varying values of testing rates for symptomatic and asymptomatic, test sensitivity, and test result reporting delay (see Table A9 in [7]), 2) self-isolation compliance for symptomatic cases and quarantine compliance for susceptible household members (see Table A10 in [7]), 3) time varying and age specific probabilities of ‘hospitalization among reported cases’ and ‘death among hospitalized’ (see Tables A2 and A3 in Additional file 1), 4) mask usage compliance (100% compliance was assumed to reduce transmission coefficient by 33% [26]), 5) contact tracing level (assumed to be at 15% based on [27], [28]), 6) percentage return to school (considred to be 50% based on [29]), 7) willingness to vaccinate (considered to be 60% based on survey results in [30], [31]), 8) variations in daily schedules during various phases of social interventions (see Table A5 in [7]), 9) percentage of asymptomatic among infected (assumed to be 35% [32]), and 10) vaccine efficacy (95% [3]).

On the first day of the simulation, the model introduced a few initial infected cases in the community and began the social mixing process. Each day the model tracked the following for each person: 1) hourly movements and locations based on their daily schedules that depend on age, employment status, prevailing social intervention orders, and quarantine/isolation status; 2) hourly contacts with other susceptible and infected; 3) vaccination status and immunity, 4) force of infection accumulation; 5) start of infection; 6) visits/consultation with a doctor (if symptomatic and insured); 7) testing (if infected and visited/consulted a doctor or asymptomatic chosen for testing either randomly or via contact tracing); 8) test reporting delay; 9) disease progression (if infected); 10) hospitalization (if infected and acutely ill); and 11) recovery or death (if infected). The AB model reports daily and cumulative values of actual infected, reported infections, hospitalized, and deaths, for each age category. A schematic diagram depicting the algorithmic sequence and parameter inputs for the AB simulation model is presented in Figure 1.

**Figure 1:**
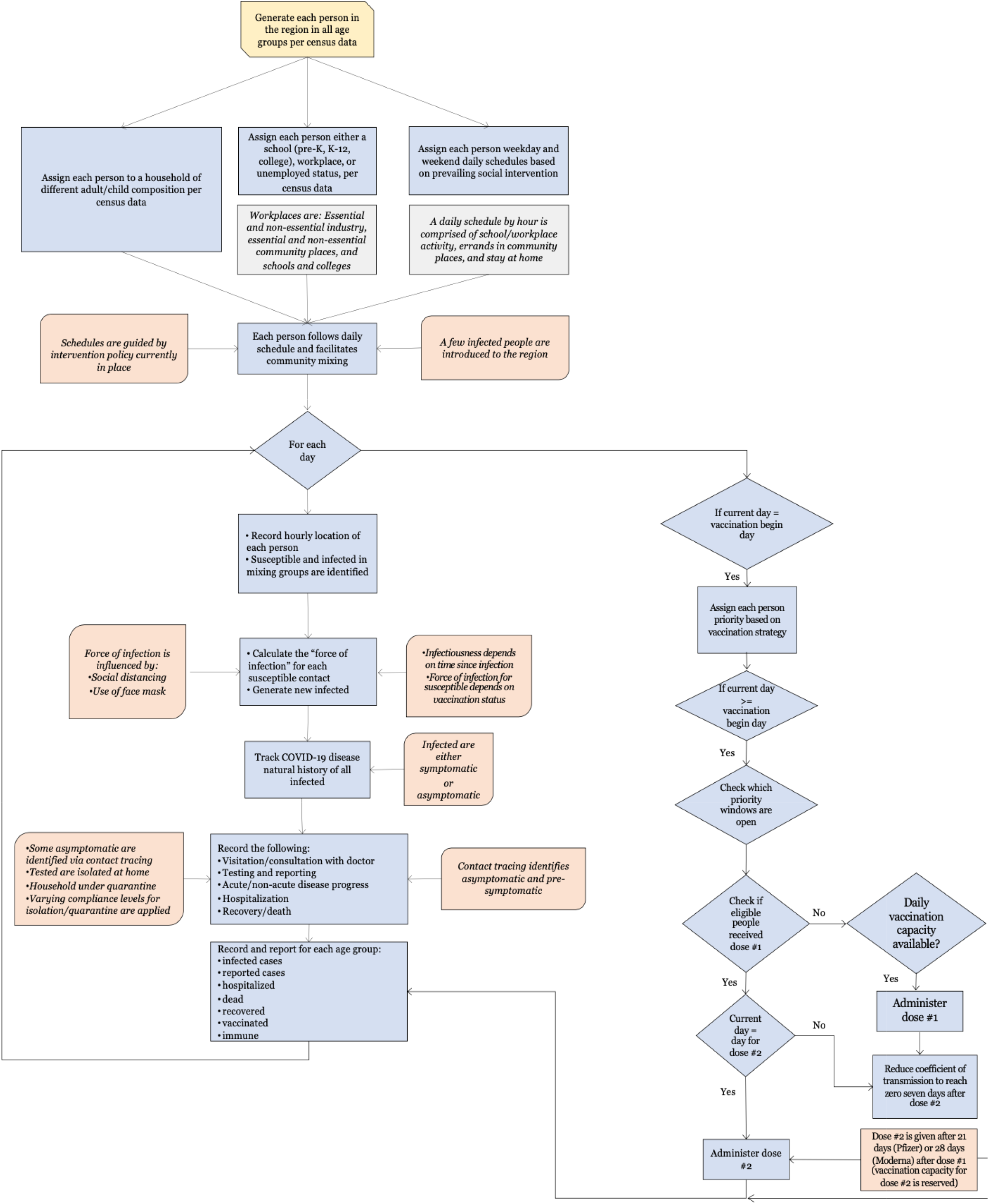
Schematic of the AB model for mimicking COVID-19 spread under social interventions and vaccination in the U.S.

For the susceptible, if vaccinated, since the exact immune response from vaccine was not known at the time of the study, we assumed a linearly increasing partial immunity for susceptible after they received the first dose, attaining full immunity after seven days after the second dose; we only considered vaccines made by Pfizer/BioNTech and Moderna, for which the second dose was administered 21 and 28 days after the first dose, respectively. The model updated the infection status of all individuals to account for new infections as well as disease progressions of infected individuals. A pseudo-code in Figure A1 in Additional file 1 depicts the major elements and structure of the AB simulation program.

For the infected, we considered the following epidemiological parameters: disease natural history with average lengths of latent, incubation, symptomatic, and recovery periods; distribution of infectiousness; percent asymptomatic; and fatality rate. The infected people were considered to follow a disease natural history as shown in Figure 2, parameters of which can be found in Table A6 of [7]. The model assumed that the recovered cases became fully immune to further COVID-19 infections. However, since this assumption was not fully supported by data at the time of the study, people recovered from COVID-19 were also considered candidates for vaccination. The duration and intensity of infectiousness was guided by a lognormal density function (see Figure 3). The function was truncated at the average length of the infectiousness period (which was considered to be 9.5 days). Asymptomatic cases were assumed to follow a similar infectiousness intensity profile but scaled by a factor, as used in the force of infection calculation (1) (see Table A4 in Additional file 1).

**Figure 2:**
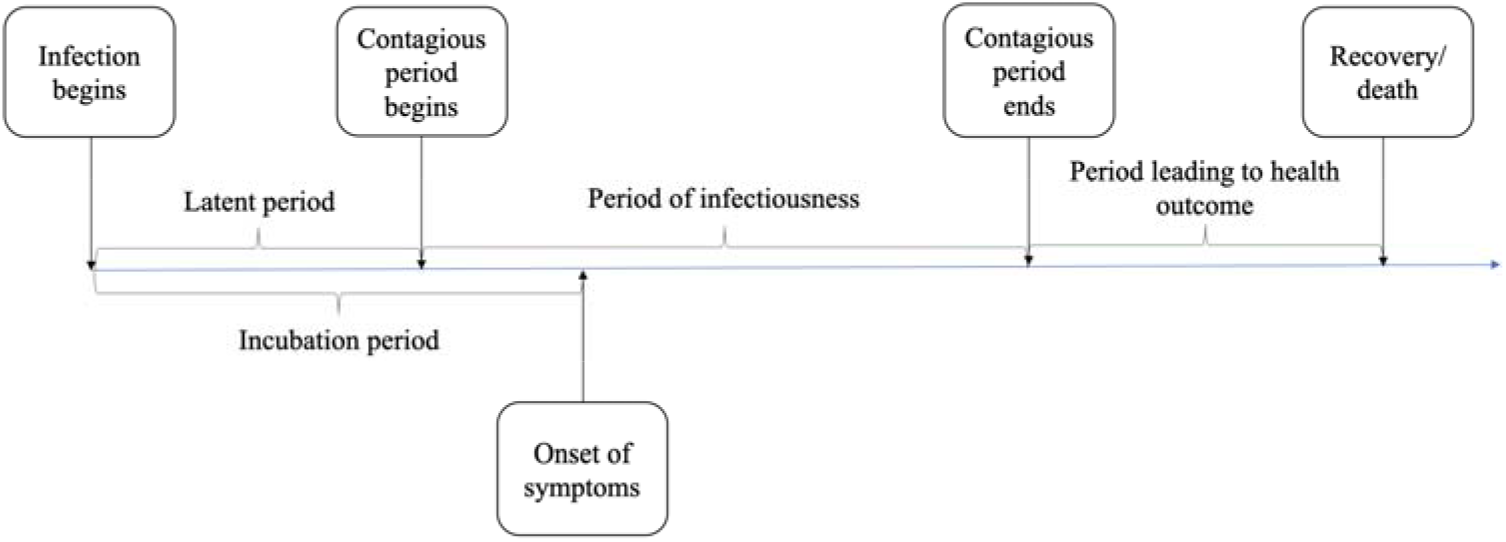
Disease natural history of COVID-19 [7]

**Figure 3:**
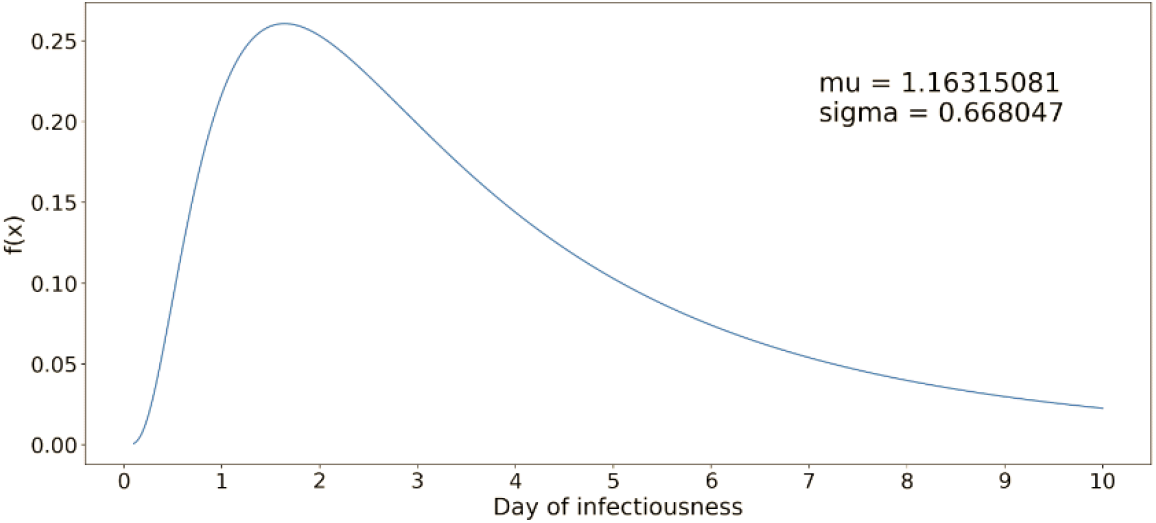
Lognormal distribution function for infectiousness profile of a COVID-19 case [7]

In what follows we describe how we compute the force of infection and used in determining the probability of infection of individuals. Each susceptible was assumed to ingest viral particles from each infected they come in contact with during the day. The total amount of ingestion of viral particles for a susceptible *i* was measured as the force of infection (*λ*_*i*_) using a modified version of the equation presented in [33]. The equation for force of infection (as presented in [33]) has three components to account for ingestions experienced from infected contacts at home, workplace/school/indoor community locations, and outdoor locations. As we had not found evidence in the literature of any significant COVID-19 transmissions at outdoor locations, we eliminated the third component and used only the first two elements of the equation as shown in (1). The first component of (1) accounts for the daily force of infection experienced by a susceptible *i* from those infected at home, and the second component accounts for workplace/school/indoor community locations.

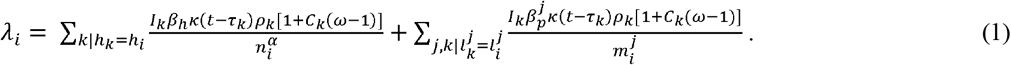

The definitions and values of parameters of (1) can be found in Table A4 in Additional file 1. The daily force of infection was considered to accumulate. However, it was assumed that if a susceptible does not gather any additional force of infection (i.e., does not come in contact with any infected) for two consecutive days, the cumulative force of infection for the susceptible reduces to zero. At the end of each day, the cumulative value of *λ*_*i*_ was used to calculate the probability of infection for susceptible *i* as 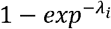 This probability was used to classify a susceptible individual as infected in the simulation model.

As observed in [7], though it was implemented for a specific region, our model is quite general in its usability for other urban regions with similar demography, societal characteristics, and intervention measures. In our model, demographic inputs (age and household distribution, number of schools for various age groups, and number of workplaces of various types and sizes) were curated from both national and local census records. Social interventions vary from region to region and the related parameters can be easily updated. Similarly, the data related to epidemiology of COVID-19 were unlikely to significantly vary from one region to another, though some adjustments of these based on population demographics may be needed.

### Model Calibration

The AB model utilized a large number of parameters, which were *demographic, epidemiological*, and *social intervention parameters*. We kept almost all of the above parameters fixed at their respective chosen values and calibrated the model by changing values for only a few. The calibrated parameters include the transmission coefficients used in calculating force of infection at home, work, school, and community places (*β*_*h*_ and 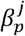) (see Table A4 in Additional file 1). The choice of the values of transmission coefficients was initially guided by [34] and thereafter adjusted at different points in time during the calibration period (until December 30, 2020). The only other parameters that were calibrated are the number of errands in the daily schedules under various intervention conditions and the percentage of workers in essential (e.g., healthcare, utility services, and grocery stores) and non-essential (e.g., offices and restaurants) workplaces who physically reported to work during different intervention periods (see Table A1 in Additional file 1). Calibration of the above parameters was done so that the daily cumulative numbers of reported infected cases from the AB simulation model closely matched the values published in the Florida COVID-19 dashboard until December 30, 2020. Figure 4 shows the cumulative values (with 95% confidence intervals) as well as the daily numbers of the reported infected cases, hospitalizations, and deaths as obtained from the simulation model. The dotted lines represent the actual numbers reported in the Florida COVID-19 dashboard for Miami-Dade County [35], the trend of which was closely followed by the numbers obtained from our AB simulation model. The actual numbers reported by the Florida COVID-19 dashboard showed large variations, which was due to reporting delays.

**Figure 4:**
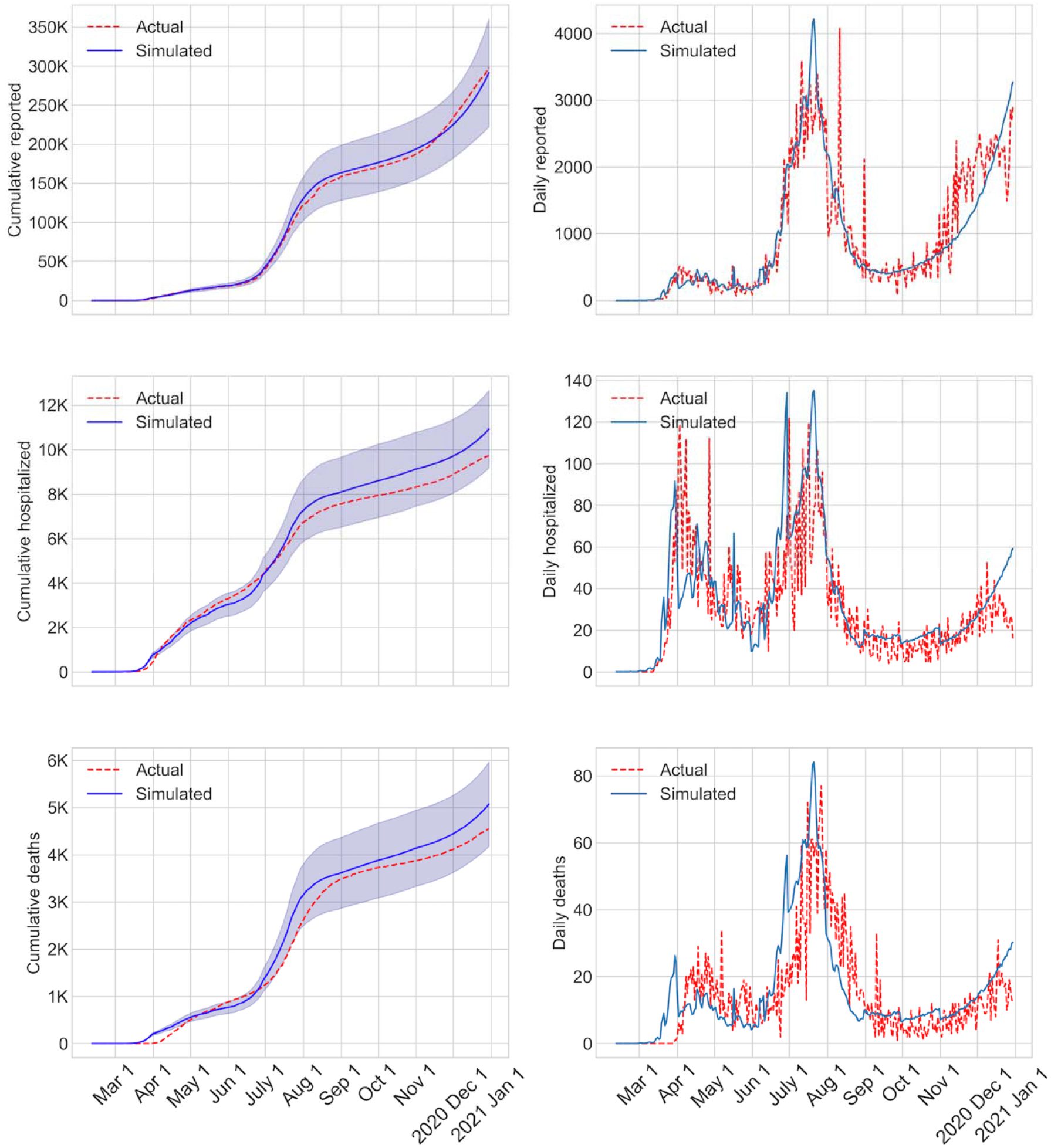
Validation graphs with cumulative and daily values of infected, reported, and deaths calibrated until Dec 30, 2020

## VACCINE PRIORITIZATION STRATEGIES

We used our AB model to examine the expected benefits of the ongoing vaccination in the U.S. using the limited supply of two types of vaccines developed by Pfizer/BioNTech and Moderna, which had the emergency approvals for distribution from the USFDA. We considered the number of vaccine doses that the two companies were contracted by the U.S. government to supply, which include the initial contracts for 100 million doses from each company and the more recent contract for an additional 100 million doses from Pfizer/BioNTech. This amounted to a total of 300 million doses, which could inoculate 150 million people with two required doses. To our knowledge, the total supply was being apportioned among the states and the counties depending on the population. Florida had approximately 6.5% of the U.S. population and the Miami Dade County had 13% of Florida’s population. Hence, we assumed that Miami Dade County would receive approximately 2.54 million doses and would be able to vaccinate 1.27 million people out of the total 2.8 million population. We also assumed that the vaccine deliveries will occur in batches starting in late December 2020 till late June 2021. Our study goal was to first determine the extent of reduction in the number of infections, hospitalizations, and deaths that we can expect to realize from the vaccination process in comparison with if no vaccines were available. Thereafter, we conducted a comparative study between three different vaccination priority schemes to determine if the outcomes (number of reported cases, hospitalized, and deaths) from those were statistically significant.

The priority strategies that were examined are broadly described here; a more complete description is presented in Figure 5. In the absence of a declared timeline for transition of eligibility from one priority group to the next, we assumed 30 days between transition. This period was extended to allow all eligible and willing to be vaccinated when the phased vaccine supply fell short of the number of people in the eligible priority group. The first strategy that we implemented was a minor ***variant of the CDC recommended strategy***: Priority 1: healthcare providers and nursing home residents; Priority 2: first responders, educators, and people of ages 75 and over; Priority 3: people of ages 65 to 74; Priority 4: people of ages 16 to 64. The CDC recommended strategy also included in priority 3 people of ages 16 to 64 with specific health conditions. Since we did not track health conditions in our AB model, we limited our priority 3 to people of ages 65 and above only. The second strategy that we implemented was an ***age-stratified strategy***: Priority 1: healthcare providers and nursing home residents; Priority 2: people of ages 65 and over; Priority 3: people of ages 55 to 64; Priority 4: people of ages 45 to 54; Priority 5: people of ages 16 to 44. The third strategy that we implemented was a ***random strategy***: Priority 1: healthcare providers and nursing home residents; Priority 2: all people of ages 16 and over. People with prior COVID-19 history were not excluded and 60% of the people were considered willing to vaccinate [30], [31].

**Figure 5:**
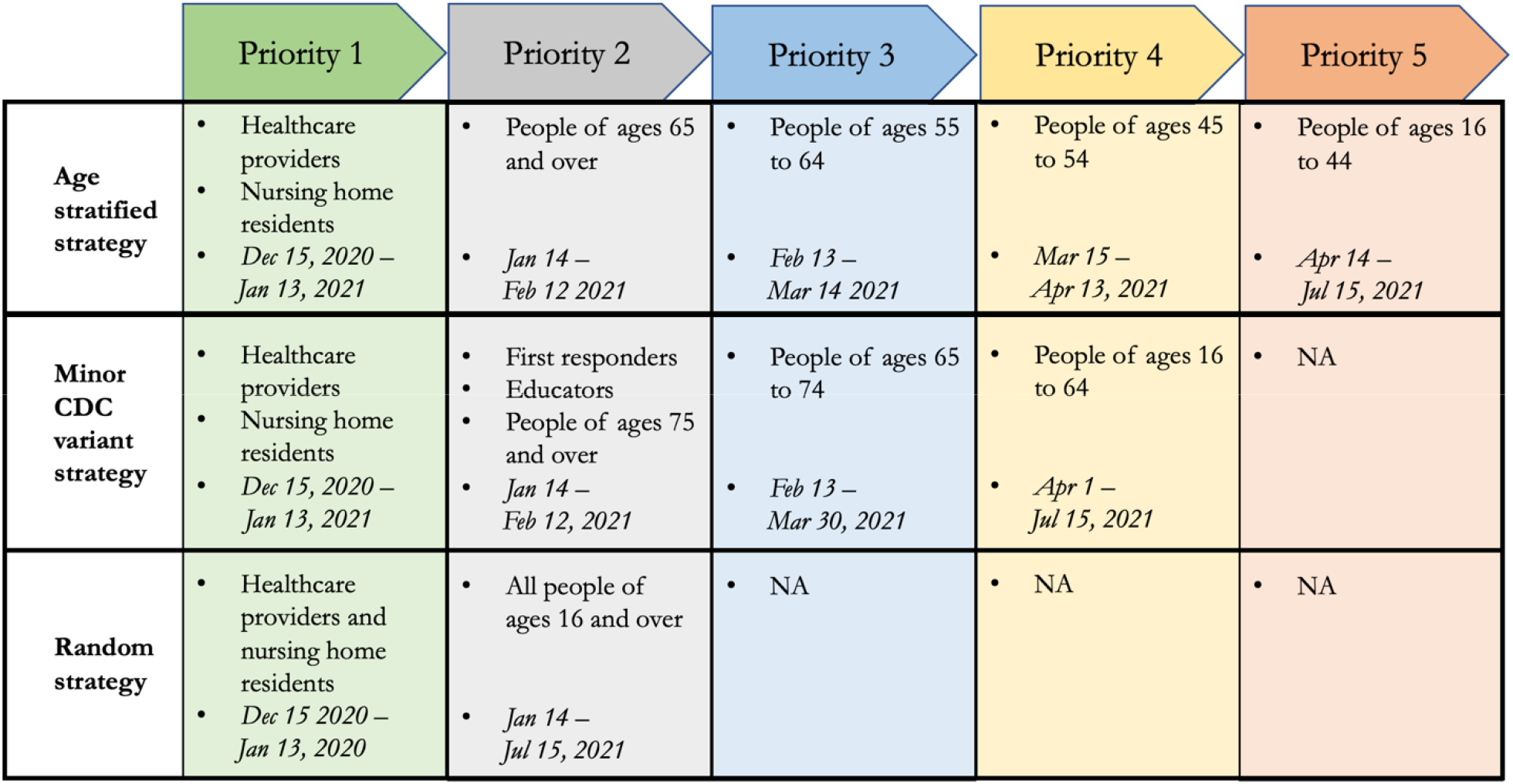
Vaccine prioritization strategies examined using AB simulation model for COVID-19 in the U.S.

## RESULTS

Figure 6 shows the plots indicating the growth over time in the number of actual reported vaccinations together with those from the three vaccination prioritization strategies that we had implemented. The vaccination began on December 15, 2020. As evident from the figure, the vaccination growth of the CDC variant strategy aligned closely with the acutal reported numbers. In the random strategy, the growth in vaccination occured faster as this strategy opened eligibility to all ages 16 and above sooner than other strategies. The age dependent strategy further staggers the eligibility and hence the vaccination grew slower than the CDC variant strategy. The flattening of the vaccination growth curves was representative of the limited vaccine supply that was considered in our simulation. The continued growth of the acutal reported vaccinations (red dotted line) was indicative of the increase in vaccine supplies since the time of our model implementation in December 2020.

**Figure 6:**
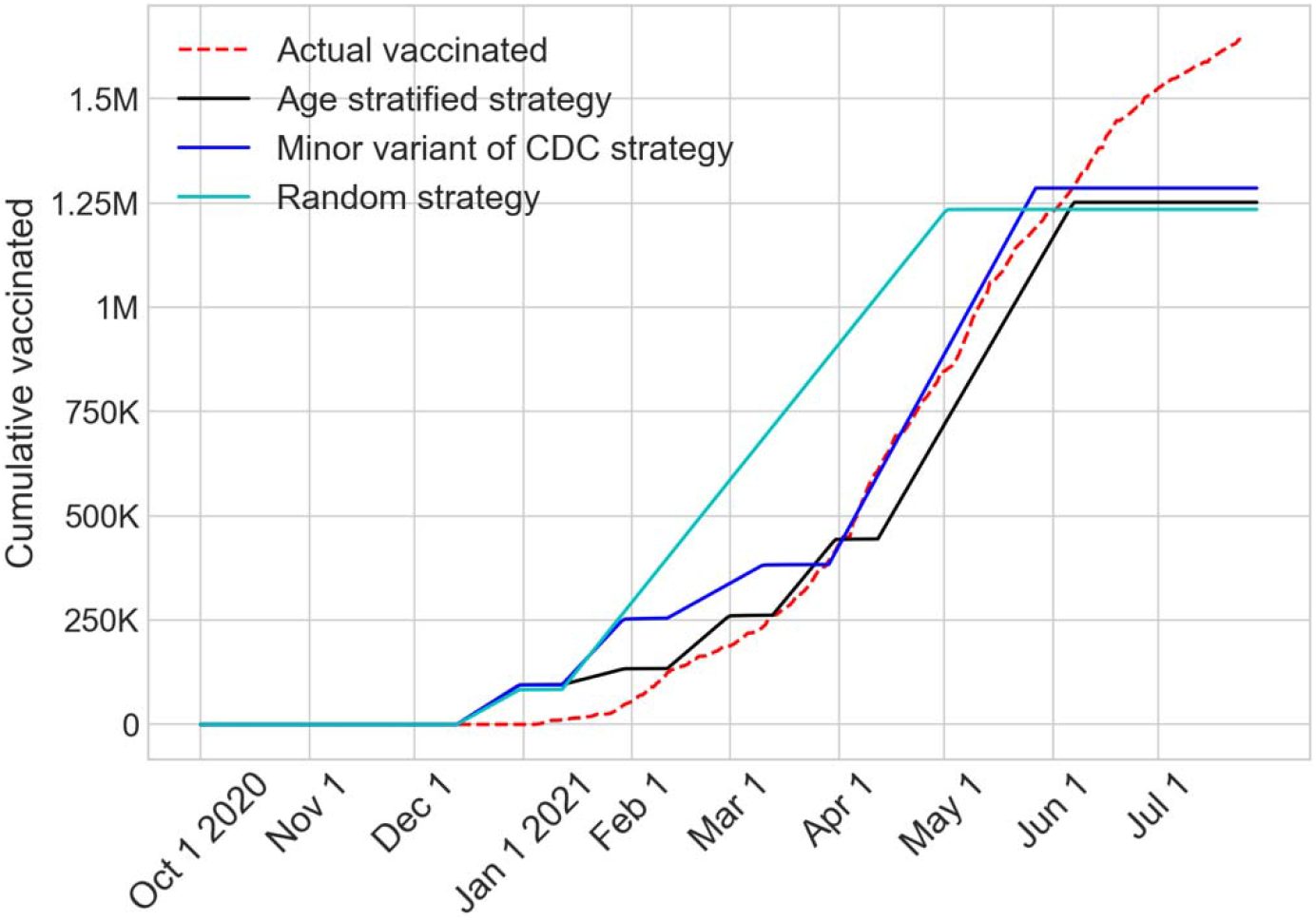
Cumulative values of actual reported and simulated vaccinations until July 31, 2021

Figure 7 shows the trends of the cumulative numbers of infected cases, reported cases, hospitalized, and deaths from the three vaccine prioritization strategies together with the no vaccination scenario until July 31, 2021 (last day of simulation). Since the confidence intervals for the cumulative values of the strategies overlap significantly, for the purpose of comparison, we chose to focus on the cumulative values on the last day of simulation. These are presented in Table 2, which summarizes the values and their 95% confidence intervals. The table also provides the time frame when the pandemic, according to the model, subsides with the new reported cases falling below the threshold of 100. We note that, the reported cases presented in Figure 7 and Table 2 were obtained directly from the simulation model. However, the hospitalization and death numbers were calculated by applying the time-varying and age-specific probabilities (as presented in Tables A2 and A3 in Additional file 1) on the reported cases.

**Table 2:** Summary of expected cumulative values (with 95% confidence intervals) on July 31, 2021 obtained by the AB model for the vaccine prioritization strategies

| Outcome \ Prioritization Strategy | Infected Cases | Reported Cases | Hospitalized | Deaths | Date when new reported cases fell below 100 |
| --- | --- | --- | --- | --- | --- |
| No vaccination | 1.71M<br>(1.66M – 1.76M) | 732K<br>(695K – 770K) | 18.7K<br>(18.3K – 19.1K) | 9K<br>(8.8K – 9.3K) | June 12, 2021 |
| Minor variant of CDC | 1.55M<br>(1.38M – 1.73M) | 659K<br>(567K – 752K) | 17K<br>(15.7k – 18.3k) | 8K<br>(7.2K – 8.8K) | May 17, 2021 |
| Age stratified | 1.58M<br>(1.43M –<br>1.73M) | 672K<br>(589K –<br>754K) | 17.6K<br>(16.6k –<br>18.5k) | 8.5K<br>(7.9k – 8.9k) | May 22, 2021 |
| Random | 1.54M<br>(1.32M –<br>1.75M) | 649K<br>(538K –<br>761K) | 17.3K<br>(15.9K –<br>18.7K) | 8.4K<br>(7.6K –<br>9.1K) | May 7, 2021 |

**Figure 7:**
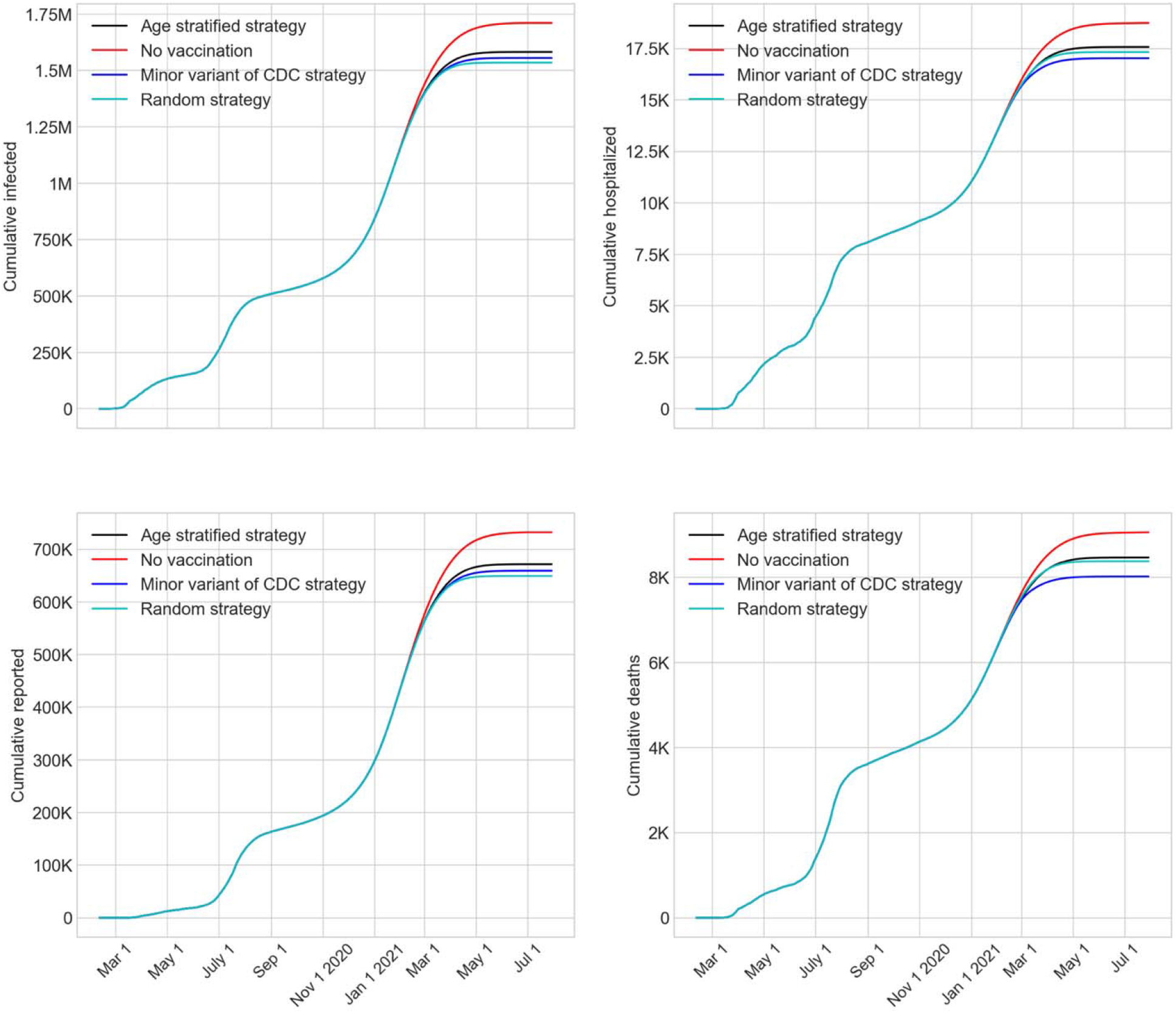
Impact of vaccination strategies on cumulative numbers of infected, reported, hospitalized, and deaths from COVID-19 until July 31, 2021

Since the performance of the three vaccine prioritization strategies, as shown in Figure 7, appeared to be similar, we conducted simple pairwise statistical comparisons of the numbers of reported cases (from column 3 of Table 2) using a test of hypothesis. According to the test results, the variant of the CDC strategy produced a statistically significantly lower number of reported cases than no vaccination (p-value 0.0204). Similar results were found for age-stratified and random strategies when compared to no vaccination. However, a comparison of the reported cases among the three vaccine prioritization strategies showed no significant statistical difference (p-values near 0.4). Pairwise comparison of the hospitalized and deaths (in columns 4 and 5 of Table 2) showed that the CDC variant strategy produced statistically significantly lower numbers compared to no vaccination (p-values 0.0014 and 0.0015, respectively). However, similar to the reported cases, the three vaccination strategies did not statistically differ among themselves in terms of hospitalizations and deaths.

The numbers from Table 2 also indicate that the CDC variant strategy achieved a reduction of 9%, 10%, 9%, and 11% for total infected, reported cases, hospitalized, and deaths, respectively, compared to the outcomes with no vaccination. Moreover, the CDC variant resulted in the pandemic subsiding below a small threshold of 100 new reported cases about a month sooner when compared with no vaccination. The CDC variant strategy also spared 5.6% of the population form being infected. The above seemingly low impact of vaccination may be attributed to the explosive growth of new reported cases that occurred in the winter months (Nov. 2020-Mar. 2021), which likely had significantly reduced the pool of susceptible people.

Figure 8 offers a further visual comparison of the impact of vaccination strategies on the percentage of population infected and the total number of deaths. A few interesting observations can be made from the figure as follows. The random strategy yielded the lowest percentage of population infected (even lower than CDC variant) as it made the age groups most active in social mixing eligible for vaccination sooner than all other strategies. For the same reason however, the random strategy yielded more deaths than CDC variant as it caused a delay in vaccination of the most vulernable elderly population who were not exclusively prioritized. Interstingly, the random strategy also performed better than the age stratified strategy in both measures of infected and deaths.

**Figure 8:**
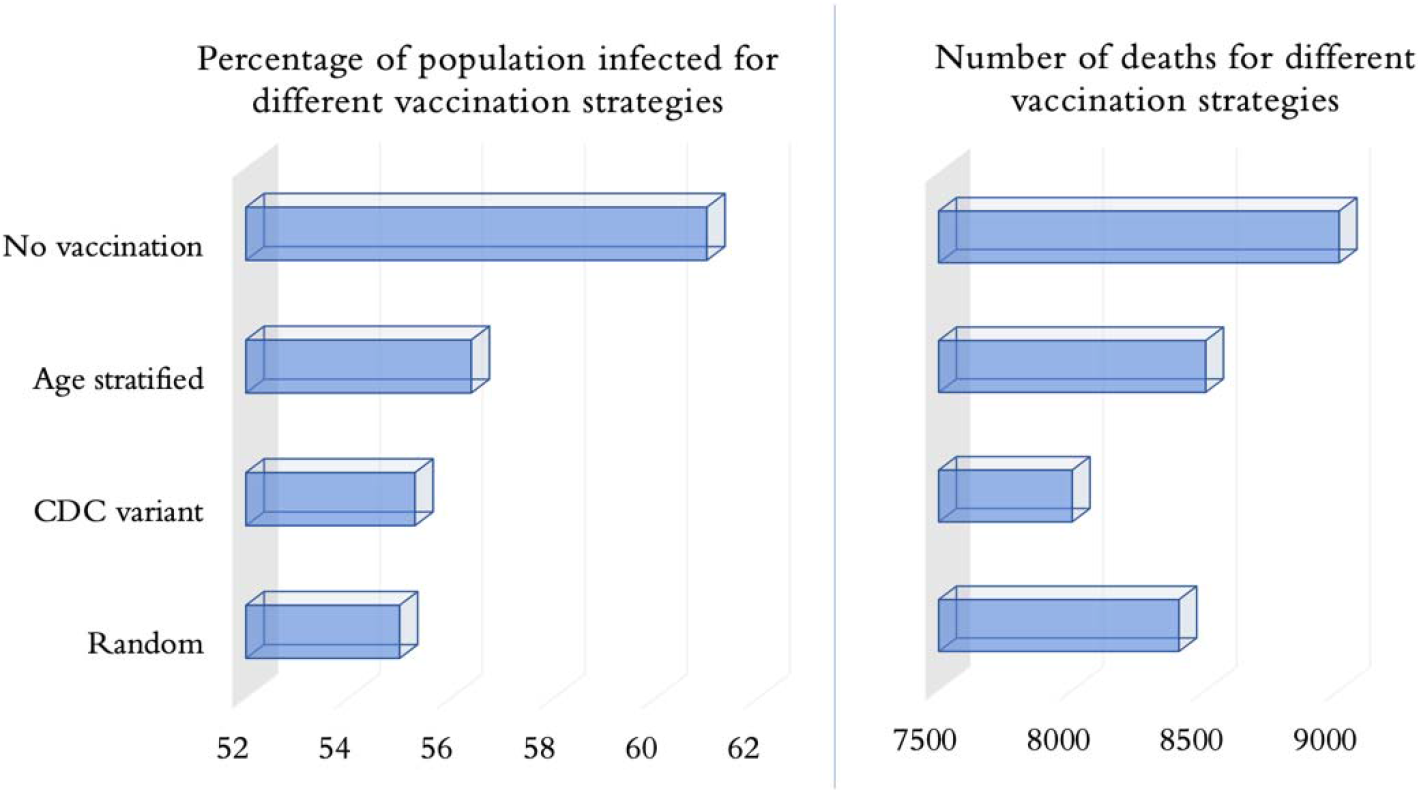
Visualization of the impacts of vaccination strategies on the percentage of total population infected an the total numbers of deaths on July 31, 2021

## CONCLUDING REMARKS

We have developed a detailed agent-based simulation model for mimicking the spread of COVID-19 in an urban region (Miami-Dade County, Florida) of the U.S. The model was calibrated using transmission coefficients and parameters guiding the daily schedules of people, and was validated using the actual reported cases from the Florida COVID-19 dashboard till December 30, 2020 (see Figure 4). On this validated model, we incorporated the vaccination process that started in the U.S. on December 15, 2020 using two different vaccines developed by Pfizer/BioNTech and Moderna. Based on the government contracts at the time of our study, we assumed availability of an estimated 2.54 million doses for Miami-Dade County to inoculate 1.27 million people (among the total population of 2.8 million) on a 2 dose regimen.

Model results indicated that the use of the available vaccines reduced the spread of the virus and helped the pandemic to subside below a small threshold of 100 daily new cases by mid-May 2021, approximately a month sooner than if no vaccines were available. Also, the vaccination was shown to reduce the number of infections by approximately 10% compared to no vaccination, which translates to sparing 5.6% of the total population from being infected. We note that, even though the vaccines were developed and approved for human use at a much faster rate than ever accomplished before, the accelerated growth of the infections, especially with the onset of the winter in the northern hemisphere, reduced the expected benefits of vaccination.

Another noteworthy finding of this study was that there were no statistical differences between the numbers of reported cases resulting from different vaccination prioritization strategies that were tested. This information should give more latitude to decision makers in urban regions across the world for distribution of the limited supply of COVID-19 vaccine. Our results suggested that instead of adhering strictly to a sequential prioritizing strategy, focus should be on distributing the vaccines among all eligible as quickly as possible.

Though our AB model is well suited to study future progression of COVID-19 and other similar respiratory type viruses, it has some limitations. The simulation model is highly granular, which while being a strength presents a challenge of appropriately estimating the wide array of its input parameters. Though we have attempted to address this challenge by conducting a sensitivity analysis on some of the critical parameters, such as levels of face mask usage, contact tracing, societal closure, and school reopening, this analysis could be extended to many other parameters. As mentioned under vaccination strategies, our model did not include health conditions that were relevant to COVID-19 (like pulmonary disease, obesity, heart problems) as attributes for people. Hence, we were not able to implement one element of the CDC recommended prioritization strategy that recommends people aged 16-64 years with underlying medical conditions to be considered in priority 3 (see Figure 5). Also, we did not consider any vaccine wastage due to complexities associated with refrigeration, distribution, and human error. We also assumed that vaccination of all priority groups occured uniformly over the eligibility periods considered, which may not reflect the reality. Also, at the time of the study, there was little available literature on the rate of immunity growth each day from the two dose vaccines, therefore, we assumed a linear growth starting with the first dose and culminating (full immunity) seven days after the second dose. Moreover, the model did not consider, the emergence of new strains of virus as the pandemic progresses, and the lower level of immunity offered by the vaccine for breakthrough infection. Finally, changes in the virus behavior might impact the comparative outcomes of different vaccine prioritization strategies, as presented in this paper.

At the time of the publication of our study, vaccine availability is still limited in many parts of the world. Our results will provide useful information for healthcare policy makers in judiciously allocating their COVID-19 vaccine supply among their population. We also believe that our findings on vaccine prioritization strategies will serve as a resource for the decision makers for future outbreaks of similar respiratory viruses. Finally, as only a limited number of studies examining vaccine prioritization strategies have been presented to the open literature, our research makes a significant addition.

## Data Availability

The datasets used and/or analysed during the current study are available from the corresponding author on reasonable request.

## LIST OF ABBREVIATIONS

AB: Agent-based
SEIR: Susceptible – exposed – infected – recovered/removed
SARS-CoV-2: Severe acute respiratory syndrome coronavirus 2
COVID-19: Coronavirus Disease 2019
USFDA: United States Food and Drug Administration
CDC: Centers for Disease Control
NAESM: NationalAcademy of Sciences, Engineering, and Medicine
QALY: Quality of Life Years
WHO: World Health Organization

## DECLARATIONS

### Ethics approval and consent to participate

Individual human data was not used in our study. Only aggregate data made available in Florida COVID-19 Dashboard was used.

### Consent for publication

Not applicable

### Authors’ contributions

Hanisha Tatapudi: Conceived and designed the model, Selection of model input parameters and data gathering, Coding and testing of the model, Design and perform the experiments, Output analysis and review, Manuscript preparation and review

Rachita Das: Selection of model input parameters and data gathering, Output analysis and review, Manuscript preparation and review

Tapas K Das: Conceived and designed the model, Selection of model input parameters and data gathering, Coding and testing of the model, Design and perform the experiments, Output analysis and review, Manuscript preparation and review

## Acknowledgements

Not applicable

## Authors’ information

Corresponding author:

Hanisha Tatapudi – corresponding author

Telephone: +1 (813) 453-3577

Rachita Das

Telephone: +1 (813) 527-1133

Tapas K. Das

Telephone: +1 (813) 843-0285

