## Supplementary information for "Impact of Vaccine Prioritization Strategies on Mitigating COVID-19: An Agent-Based Simulation Study using an Urban Region in the United States"

Appendix

**Table A1:** Social mixing parameters used in model calibration

| Social interventions | % of essential workers reporting to work | % of non-essential community workers reporting to work | % of non-essential industry workers reporting to work | Number of errands in daily schedule |
| --- | --- | --- | --- | --- |
| Before lockdown  (stay at home order) | 100% | 100% | 100% | 3/1/3^1^ |
| During lockdown  (starting March 17, 2020) | 50% | 0% | 0% | 0.4 |
| Phase I reopening  (starting May 18, 2020) | 70% | 60% | 25% | 0.8 |
| Phase II reopening  (starting June 5, 2020) | 70% | 70% | 60% | 1.2 |
| Phase III reopening  (Sep 25, 2020) | 70% | 30% | 20% | 2 |

^1^ 3 weekday errands for unemployed / 1 weekday errand for employed / 3 weekend errands for all.

**Table A2:** Time varying and age-specific probability of hospitalization among reported cases^1^ from March – October 2020

| Age groups | March | April | May | June | July | August | September | October |
| --- | --- | --- | --- | --- | --- | --- | --- | --- |
| 0-4 | 0 | 0.05 | 0.15 | 0.04 | 0.02 | 0.03 | 0.02 | 0.03 |
| 5-14 | 0 | 0.05 | 0.04 | 0.01 | 0.01 | 0.01 | 0.01 | 0.01 |
| 15-24 | 0.05 | 0.03 | 0.04 | 0.01 | 0.01 | 0.01 | 0.01 | 0.01 |
| 25-34 | 0.06 | 0.05 | 0.05 | 0.03 | 0.01 | 0.01 | 0.01 | 0.01 |
| 35-44 | 0.13 | 0.09 | 0.06 | 0.04 | 0.01 | 0.02 | 0.02 | 0.01 |
| 45-54 | 0.17 | 0.13 | 0.08 | 0.05 | 0.02 | 0.02 | 0.03 | 0.02 |
| 55-64 | 0.22 | 0.2 | 0.17 | 0.09 | 0.04 | 0.03 | 0.05 | 0.03 |
| 65-74 | 0.4 | 0.35 | 0.32 | 0.14 | 0.07 | 0.04 | 0.07 | 0.07 |
| 75-84 | 0.55 | 0.49 | 0.48 | 0.18 | 0.11 | 0.08 | 0.15 | 0.1 |
| 85-100 | 0.57 | 0.61 | 0.58 | 0.25 | 0.17 | 0.1 | 0.18 | 0.13 |

^1^ Probabilities derived from data reported by the Florida COVID-19 Dashboard for Miami-Dade county [35].

**Table A3:** Time varying and age-specific probability of death among hospitalized cases^1,2^ from March – October 2020

| Age groups | March | April | May | June | July | August | September | October |
| --- | --- | --- | --- | --- | --- | --- | --- | --- |
| 0-4 | 0 | 0 | 0 | 0 | 0 | 0 | 0 | 0 |
| 5-14 | 0 | 0 | 0 | 0 | 0.06 | 0 | 0 | 0 |
| 15-24 | 0 | 0 | 0.05 | 0 | 0.05 | 0.03 | 0 | 0 |
| 25-34 | 0 | 0.04 | 0.02 | 0.02 | 0.07 | 0.13 | 0 | 0 |
| 35-44 | 0.04 | 0.06 | 0.09 | 0.05 | 0.13 | 0.07 | 0.09 | 0.03 |
| 45-54 | 0.09 | 0.1 | 0.07 | 0.17 | 0.22 | 0.15 | 0.1 | 0.14 |
| 55-64 | 0.2 | 0.17 | 0.17 | 0.26 | 0.44 | 0.45 | 0.31 | 0.3 |
| 65-74 | 0.31 | 0.33 | 0.25 | 0.58 | 0.84 | 0.97 | 0.61 | 0.5 |
| 75-84 | 0.37 | 0.4 | 0.46 | 0.85 | 1 | 1 | 0.85 | 0.79 |
| 85-100 | 0.77 | 0.61 | 0.59 | 1 | 1 | 1 | 1 | 1 |

^1^ Probabilities derived from data reported by the Florida COVID-19 Dashboard for Miami-Dade county [35].

^2^ Deaths reported by the dashboard are total daily numbers including those not hospitalized. However, only a small percentage of the deaths occurred outside hospitals, hence, our probability estimates for death among hospitalized cases are slightly higher.

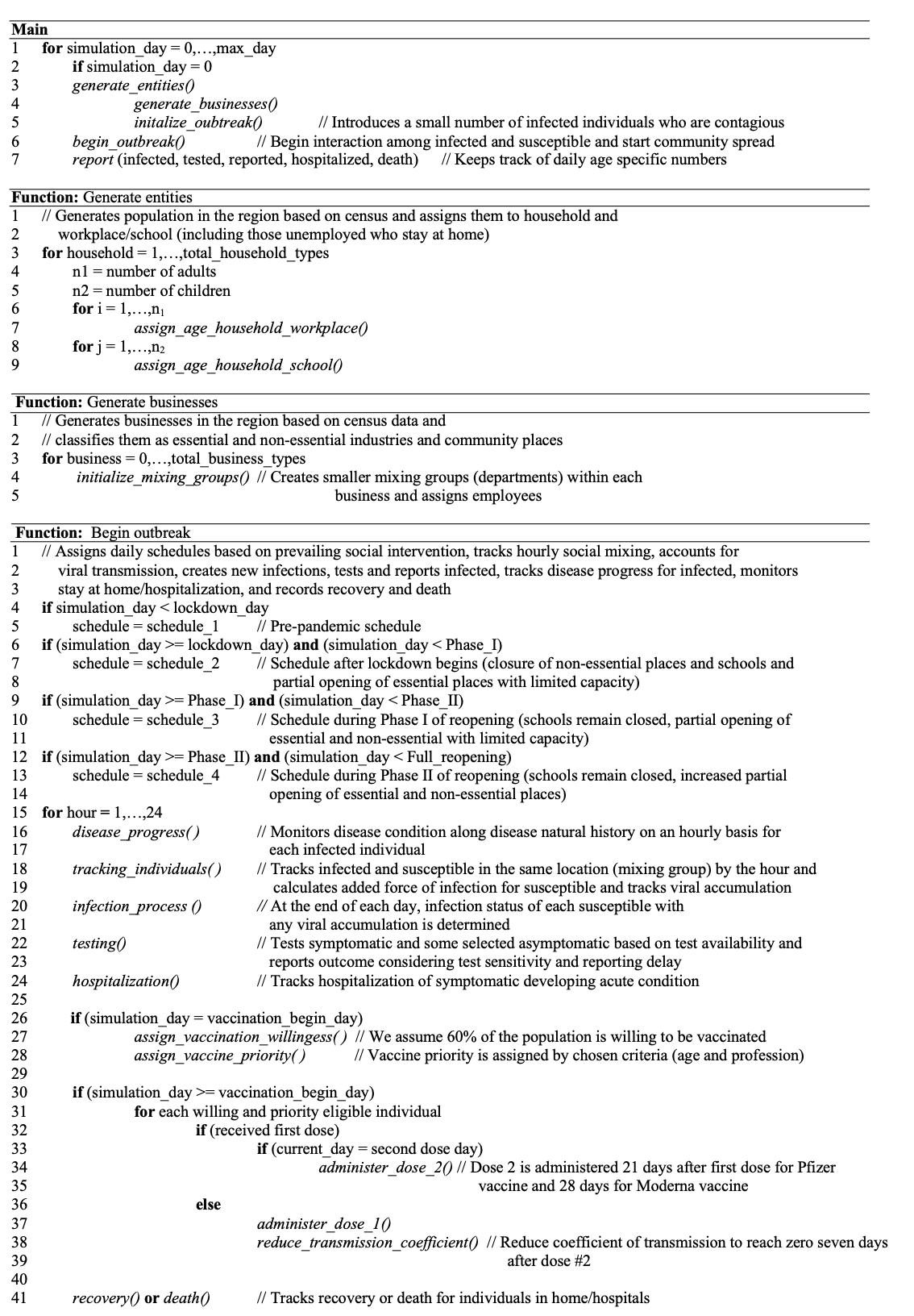

**Figure A1:** Pseudo-code for agent-based simulation model of COVID-19 with implementation of two-dose vaccines

**Table A4:** Parameters of the force of infection (equation (1))

| **Parameter** | **Description** | **Value** | | | |
| --- | --- | --- | --- | --- | --- |
| $I_{k}$ | Infected status of an individual $k$ | 1 if infected and 0 otherwise | | | |
| $\beta_{h}$ | Transmission coefficient at home**^1,2^** |  | **Period** | | |
|  |  | 0.1 | Before stay-at-home orders | | |
|  |  | 0.1 | During stay-at-home orders | | |
|  |  | 0.1 | During Phase I | | |
|  |  | 0.1 | During Phase II | | |
|  |  | 0.1 | After face mask implementation | | |
| $\beta_{p}^{j}$ | Transmission coefficient at school, work, and community places**^1,2^** | **School** | **Work** | **Community** | **Period** |
|  |  | 0.025 | 0.5 | 0.275 | Before stay-at- home order |
|  |  | - | 0.35 | 0.44 | During stay-at- home order |
|  |  | - | 0.35 | 0.37 | During Phase I |
|  |  | - | 0.5 | 0.75 | During Phase II |
|  |  | - | 0.165 | 0.2475 | Phase II with facemask^5^ |
|  |  | 0.33 | 0.165 | 0.2475 | Phase III and school reopening^6^ |
| $\kappa(t)$ | **^3^**Infectiousness at time *t* (*t* denotes the elapsed time after completion of latency) | Lognormal function value at *t* with $\mu= 1.16315081$ and $\sigma= 0.668047$ | | | |
| $\rho_{k}$ | **^4^**Relative infectiousness of individual k | 1 | | | |
| $C_{k}$ | **^4^**Scaling factor for mild/asymptomatic vs severe infection | 1 if severe/symptomatic, 0 for mild/asymptomatic | | | |
| $\omega$ | **^4^**Scaling factor for infectiousness for a mild vs severe infection | 2 for severe infection relative to a mild one | | | |
| $\alpha$ | **^4^**Scaling factor for household size | 0.8 | | | |
| $n_{i}$ | Number of people in the household of individual $i$ | Calculated from simulation | | | |
| $m_{i}^{j}$ | Number of people in the place type *j* where individual $i$is | Calculated from simulation | | | |

**^1^** Choice of these parameters were guided by Ferguson [34], literature estimates of R0 for SARS-CoV-2, and prevailing interventions. Transmission coefficients were subsequently calibrated to arrive at the values given here.

**^2^** In model implementation, all transmission coefficients were multiplied with age-dependent scaling factors of 0.25, 1, and 3 for age groups 0-14, 15-84, 85 and above, respectively. This was done to account for widely varying infection rates across age groups.

**^3^** Parameters of the lognormal distribution function were selected to have the mean length of infection as 4 days and a standard deviation of 3 days [36].

**^4^** Selected from Ferguson [34].

**^5^** Face masks reduce transmission coefficient by 33% with 100% compliance [26].

**^6^** Transmission coeffecient at school is assumed to be two times the transmission coefficient at workplaces [25].
